# Network representations of digital clock drawings for cognitive impairment detection

**DOI:** 10.64898/2026.10.15.26365699

**Authors:** Adlin Pinheiro, Huimin Cheng, Cody Karjadi, Katherine Gifford, Serkalem Demissie, Kathryn L. Lunetta, Chunyu Liu, Rhoda Au, Shariq Mohammed

## Abstract

**INTRODUCTION:** The digital clock drawing test (dCDT) is a cognitive screening tool capturing high-resolution pen-movement data. Existing approaches for classifying cognitive impairment from dCDTs rely on selected features or image-based methods. We propose representing dCDTs as structural networks to classify cognitive impairment and characterize spatial organization in clock drawings.

**METHODS:** Weighted networks were constructed from 3,362 Framingham Heart Study participants, with and without cognitive impairment. Network representations were generated separately for command and copy conditions, with nodes representing pre-defined clock components (digits, hands, clock face) and edges capturing pairwise spatial relationships between components. Random forest models and graph convolutional networks were trained to predict cognitive impairment from the networks. Models were benchmarked against demographics-only and established summary feature baselines.

**RESULTS:** Network-based models achieved AUC values comparable to the established summary feature baseline across command-only and copy-only conditions. Differences in spatial organization between the cognitively intact and impaired groups were greatest in the upper-right clock region, involving digits 1 and 2, the clock hands, and the surrounding clock face. Differences between the command and copy networks were larger among cognitively impaired participants.

**DISCUSSION:** The proposed network representations provide an objective, scalable, and flexible framework for representing clock drawings for use in classification models. dCDT networks matched the predictive performance of established summary feature-based approaches and identified clock regions where spatial organization differed by cognitive status. After external validation, this approach has the potential to be incorporated within a clinical workflow to support automated and interpretable cognitive screening.

## 1 Introduction

The Clock Drawing Test (CDT) is a widely used cognitive screening tool in clinical practice, requiring individuals to draw a clock from memory with the hands set to a specified time. Participants are typically asked to complete two conditions: drawing a clock from memory (command) and copying a provided clock image (copy). The digital version of the CDT (dCDT) extends this by using a digital pen to capture high resolution pen movements, recording the spatial coordinates, timing, and pressure for each point. This data provides a rich window into the interplay between executive function, visuospatial reasoning, and fine motor control, which are cognitive domains known to be affected early in the course of neurodegenerative disease.^1^ This makes the dCDT a compelling candidate for scalable, objective cognitive screening.

Several computational and machine learning frameworks have been developed to extract predictive information from dCDT recordings. The most established approach derives summary features extracted from the pen data to capture drawing efficiency, information processing speed, spatial organization, and motor characteristics, and uses them as inputs to classification models.^2–7^ Although predictive, this approach requires subjective decisions about which features to include, how to handle correlated predictors, and how to manage missing data when clock components are omitted from the drawing. A second approach applies deep learning by treating the completed drawings as images.^8–10^ This strategy bypasses subjective feature engineering but sacrifices interpretability and discards the temporal information that is unique to the digital pen.

A promising complementary direction is the representation of dCDT data as networks. A recent study used directed acyclic graphs to model interactions among graphomotor and time-based summary features from the dCDT, finding differences in directional dependencies between command and copy conditions among participants who completed error-free digital clock drawings.^11^ This work demonstrated that relational representations of dCDT data can reveal structural patterns not accessible from individual features in isolation. However, because the nodes of these networks represented summary features rather than physical clock components, the resulting network structure reflected statistical associations among derived features rather than the actual spatial geometry of the drawing.

We propose a fundamentally different and novel network representation in which nodes correspond to individual physical clock components (e.g., specific digits, the hour and minute hands, and spatial regions of the clock face) and edge weights encode the spatial proximity between those clock components as drawn by the participant. This representation preserves the full spatial geometry of the clock drawing at the component level without collapsing it into scalar summary statistics. Because each node corresponds to an actual element of the clock, the network is directly interpretable. That is, nodes with no edges reflect omitted components, edge weights reflect spatial positioning, and structural differences between networks can be localized to specific clock regions. Furthermore, because the network representation is defined by a fixed node set and a principled distance-based weighting scheme, it is fully objective and reproducible across samples without investigator-specific feature engineering.

Using dCDTs from the Framingham Heart Study (FHS), we construct individual-level network representations for both the command and copy test conditions and evaluate their ability to classify cognitive impairment using random forest models and graph convolutional networks. We evaluate classification performance against established summary features and identify where the largest group differences appear in the spatial structure of the clock drawing. Our central objectives are to (i) test whether a network-based representation can match the predictive performance of existing features that already capture rich information from the drawing process, and (ii) show that this fully objective, spatially grounded network representation offers meaningful structural and interpretive benefits.

## 2 Methods

### 2.1 Study Sample

The FHS is a longitudinal, prospective cohort study established in Framingham, MA in 1947, enrolling several generations of participants, their family members, and spouses, starting with the recruitment of the Original cohort. The children of the Original cohort participants and their spouses were then recruited in 1971, comprising the Offspring cohort. The Omni 1 cohort, recruited in 1995, consisted primarily of racial and ethnic minorities to reflect the changing demographics of Framingham, MA. In 2002, children of the Offspring cohort were recruited for the Third Generation cohort as well as a second minority cohort, Omni 2.

In 2011, the dCDT was introduced as part of a battery of tests for standard neuropsychological assessments. Participants were asked to draw a clock from memory, with the hands pointing to ten past eleven (command condition). In most cases, after a short interval, participants were also asked to copy an image of a clock that was provided to them (copy condition).

Participants were eligible for the present study if they had completed both a command and copy dCDT on the same occasion, on or before December 31, 2022. If a participant had undergone multiple dCDTs, the most recent was selected. Participants were excluded if they had a test with pen malfunctions, had missing demographic information, or had a history of stroke. The final sample comprised 3,362 participants.

### 2.2 Cognitive Impairment Adjudication

Participants can be flagged for dementia through self-report, report by a family member or physician, a drop in Mini-Mental State Examination score, or identification through an FHS ancillary study.^12^ All suspected cases of cognitive impairment in the FHS are adjudicated by a review panel consisting of at least one neurologist and one neuropsychologist. The panel confirms whether or not a participant has mild cognitive impairment (MCI) or dementia of any cause and the date of onset using all available information such as results from neuropsychological assessments, medical records, brain imaging studies, and autopsy data when available.

### 2.3 dCDT Recording and Pre-Processing

The digital pen (Anoto, Inc.) and its accompanying software record points every 12-13 milliseconds, capturing time, pressure, *x* and *y* coordinates. Each stroke is subsequently classified as originating from a particular clock component such as a digit, clock hand, or clock face using the ClockSketch software, with any classification errors manually corrected by a research assistant.^13^

A dCDT can be reconstructed by plotting the *x* and *y* coordinates of all points captured by the digital pen. For data pre-processing, the coordinates were scaled such that all points fit into a 1 × 1 unit square, preserving the aspect ratio and spatial relationships between clock components. This normalization facilitates comparison across dCDTs by removing variability due to differences in drawing size and starting position. The scaling factor was stored and included as a predictor in downstream classification models.

Next, a comprehensive set of summary features identified through a systematic review of the dCDT literature and previously shown to be predictive of cognitive outcomes were extracted from the pen data.^1,3,4,6,7,14–17^ These features have been categorized to capture drawing efficiency, information processing, spatial reasoning, and simple and complex motor features. Definitions of these summary features can be found in Supplementary Table S1.

### 2.4 Network Preliminaries

A network (graph) is a structure used to describe relationships or connections between objects. A network denoted by *G* = (*V, E*) is described by a set *V* of nodes and a set *E ⊂ V × V* of edges. A network is represented by its adjacency matrix *A* ∈ {0, 1}*^p×p^*, where *p* is the number of nodes. For an undirected and unweighted network, *A*(*i, j*) = 1 if and only if link (*i, j*) ∈ *E*. Because edges are bidirectional in an undirected network, *A*(*i, j*) = *A*(*j, i*), and *A* is symmetric with *p* real-valued eigenvalues. For a directed graph, *A* is not symmetric and its eigenvalues are complex. Networks can also have weighted edges where *A*(*i, j*) takes values in the interval [0, 1].

#### Matrix Representations

A network may also be represented by its Laplacian matrix, *L* = *D-A*, where *D* is a diagonal matrix containing the degree of each node. The degree of a node is the number of edges connected to it in an unweighted network. For a weighted network, the degree is the sum of all edge weights connected to the node. *L* is a positive semi-definite matrix with at least one zero eigenvalue.^18^ An alternative representation is the normalized Laplacian matrix, 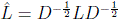 which accounts for node degree heterogeneity.

#### Graph Spectra

The collection of eigenvalues obtained from the eigendecomposition of an adjacency matrix is known as the *graph spectra*. Spectral graph parameters capture extensive information about both the global and local structure of a graph.^19^ For example, the largest eigenvalue of the adjacency matrix *A*, known as the *spectral radius*, is a dynamic mean value of node degrees, taking into account neighbors of nodes and the neighbors of the neighbors.^19^ The second smallest eigenvalue of *A* is related to the graph’s expansion parameter, where smaller values indicate that the edges are more evenly distributed throughout the graph.^20^ Additionally, the smallest eigenvalue of *A* is closely related to how bipartite a graph is.^21^ The second smallest eigenvalue of the Laplacian *L*, known as the *algebraic connectivity*, is a measure of a graph’s robustness, where higher values indicate stronger connectivity, and is equal to zero if *G* is disconnected.^22,23^ This is similarly reflected in the second smallest eigenvalue of the normalized Laplacian matrix.^24^

#### Edit Distance

A metric measuring the similarity between any two graphs is the *edit distance*. The edit distance between two networks *G*_1_ and *G*_2_ is

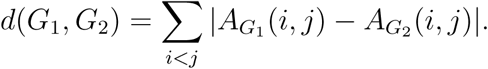

If *G*_1_ and *G*_2_ are unweighted networks with the same node set, the edit distance is the number of edge insertions or deletions required to transform *G*_1_ into *G*_2_ (or vice versa). Conversely, the edit distance for weighted networks is the sum of the absolute differences between all corresponding edge weights of *G*_1_ and *G*_2_.

### 2.5 dCDT Network Construction

Each clock drawing in the sample was represented as a weighted, undirected network. A set of 19 fixed nodes was selected to include standard clock components and additional components intentionally introduced to better capture spatial patterns and structural organization of the drawing. The selected node set is

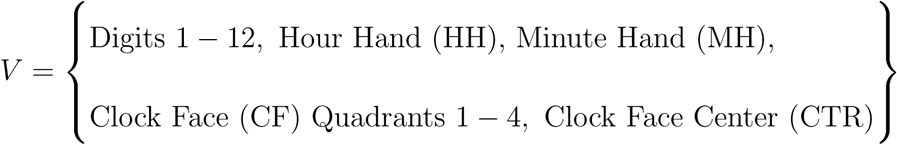

The clock face is subdivided into four quadrants, defined by angles measured clockwise from the 12:00 position (0°). Specifically, Quadrant 1 (Q1) includes angles from 0^◦^ to 90^◦^, Quadrant 2 (Q2) from 90^◦^ to 180^◦^, Quadrant 3 (Q3) from 180^◦^ to 270^◦^, and Quadrant 4 (Q4) from 270^◦^ to 360^◦^. This subdivision of the clock face allows the capture of patterns or deficits in spatial organization across different regions of the clock face.^17^ Additionally, the clock face center node provides a fixed reference point for the network representation, allowing the capture of distances of the clock elements to the center.

Any two nodes *i* and *j* are connected if and only if the corresponding clock components are drawn on the test. Therefore, a node is isolated (i.e., not connected to any other node) if the component is omitted on the dCDT. Weights are assigned to edges to reflect the spatial proximity between the clock components. The weight of the edge connecting nodes *i* and *j* is defined as *w*_(*i,j*)_ = exp(–*d*_(*i,j*)_) if (*i, j*) ∈ *E*, and 0 otherwise, where *d*_(*i,j*)_ is the standardized Euclidean distance between the centroids of clock components corresponding to nodes *i* and *j*. The centroids of convex hulls, defined as the smallest convex polygon that completely encloses a set of points, were used to identify the geometric centers of each clock component. Because edge weights depend on where each component is drawn, misplaced digits or clock hands change the corresponding edge weights. For example, a minute hand pointing toward 10 rather than 2 reduces the weight of the edge between the minute hand and digit 2. Since all distances are standardized and bounded such that 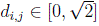, the corresponding edge weights are similarly bounded by 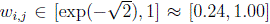. Smaller distances yield higher edge weights, reflecting stronger spatial proximity.

In the event that a clock component was drawn twice, the distances from each instance of the component to other nodes were averaged to compute the corresponding edge weights. Extraneous strokes and incorrect numbers that were not classified as one of the predefined clock components were not explicitly represented in the current 19-node network. The code used to convert digital pen data to structural networks can be found at: https://github.com/shariq-mohammed/clocknet.

### 2.6 Prediction Model Development and Evaluation

Demographics including age, sex, APOE-*€*4 status, FHS cohort, and education were obtained for each participant along with cognitive status at the time of the dCDT. The primary outcome was cognitive impairment, defined as MCI or dementia of any cause. An overview of our pipeline is provided in Figure 1.

**Figure 1:**
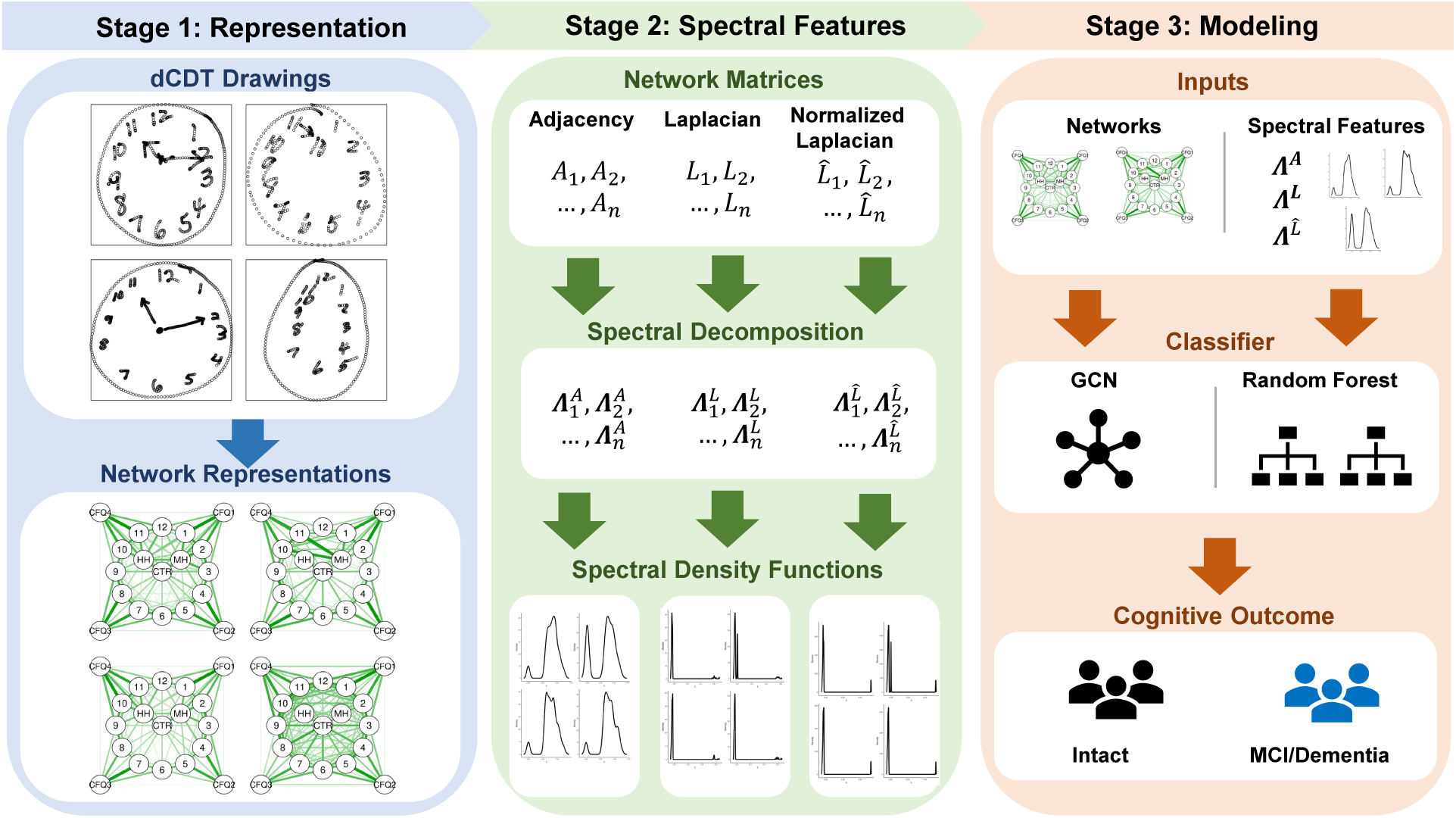
Overview of the dCDT network construction and classification pipeline. For each command and copy drawing, a weighted, undirected network was constructed with 19 nodes representing clock components (digits 1-12, hour hand, minute hand, clock face center, and four clock face quadrants) and edges encoding standardized spatial proximity between component centroids. Adjacency (*A*), Laplacian (*L*), and normalized Laplacian (*L*) matrix representations were derived and their eigenvalues computed for use as predictors in random forest models. Graph convolutional networks (GCNs) were also used as an alternative classification approach. Secondary analyses explored spectral density function representations (summarized through functional principal component analysis) and edge thresholding prior to eigenvalue computation.

#### 2.6.1 Feature Sets and Models

Since this is the first study to represent dCDTs as structural networks, we conducted a systematic evaluation of multiple network-based classification approaches to identify the most effective and informative representation. All network-based models were benchmarked against two conventional baselines: a demographics-only model and a model incorporating a comprehensive set of existing dCDT summary features. Critically, the summary feature set was not drawn from any single published study but was assembled through a thorough review of the dCDT literature, representing a complete collation of features currently available (Supplementary Table S1). This ensures that the comparison reflects the ceiling of existing feature-based approaches rather than any particular implementation, providing the most rigorous possible benchmark for evaluating the added value of network-based representations. Network-based models ranged in complexity from interpretable graph-theoretic summary measures (Supplementary Table S2) to eigenvalue-based spectral representations of different matrix forms of the network, to graph convolutional networks that operate directly on the full network structure. An overview of all models evaluated is provided in Table 1.

**Table 1:**
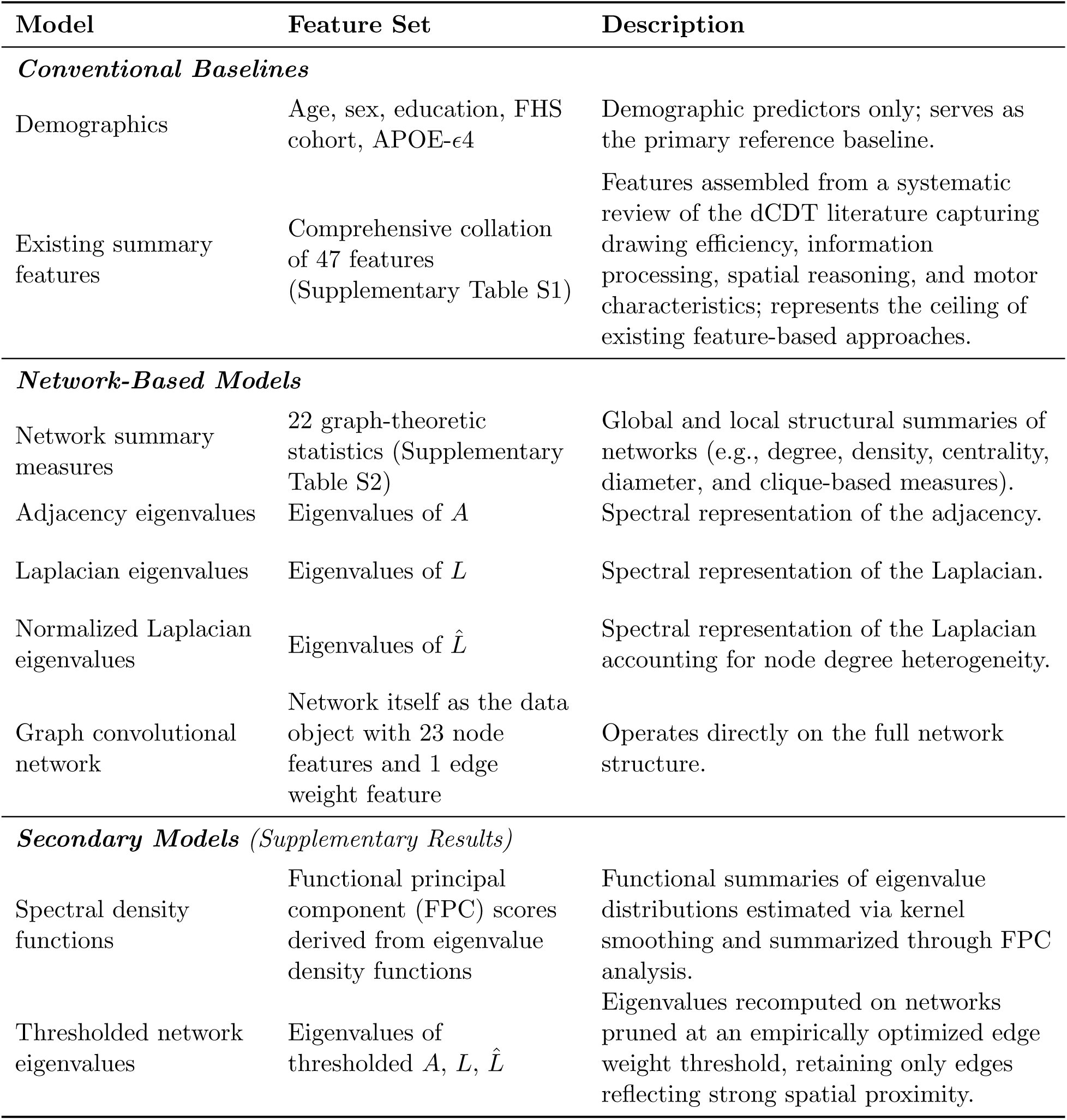
Overview of classification models for prediction of cognitive impairment from dCDT networks. All models include demographic covariates (age, sex, education, Framingham cohort, and APOE-ε4 carrier status). All models except the demographics-only baseline additionally include total time to completion, total ink time, total think time, and image scaling factor as covariates. Each network-based model was evaluated separately under command-only, copy-only, and combined command and copy conditions. Network-based predictors were derived from participant-level weighted undirected networks with a fixed 19-node set (digits 1–12, hour hand, minute hand, clock face center, and four clock face quadrants).

| Model | Feature Set | Description |
| --- | --- | --- |
| *Conventional Baselines* |  |  |
| Demographics | Age, sex, education, FHS cohort, APOE-ε4 | Demographic predictors only; serves as the primary reference baseline. |
| Existing summary features | Comprehensive collation of 47 features (Supplementary Table S1) | Features assembled from a systematic review of the dCDT literature capturing drawing efficiency, information processing, spatial reasoning, and motor characteristics; represents the ceiling of existing feature-based approaches. |
| *Network-Based Models* |  |  |
| Network summary measures | 22 graph-theoretic statistics (Supplementary Table S2) | Global and local structural summaries of networks (e.g., degree, density, centrality, diameter, and clique-based measures). |
| Adjacency eigenvalues | Eigenvalues of $A$ | Spectral representation of the adjacency. |
| Laplacian eigenvalues | Eigenvalues of $L$ | Spectral representation of the Laplacian. |
| Normalized Laplacian eigenvalues | Eigenvalues of $\hat{L}$ | Spectral representation of the Laplacian accounting for node degree heterogeneity. |
| Graph convolutional network | Network itself as the data object with 23 node features and 1 edge weight feature | Operates directly on the full network structure. |
| *Secondary Models (Supplementary Results)* |  |  |
| Spectral density functions | Functional principal component (FPC) scores derived from eigenvalue density functions | Functional summaries of eigenvalue distributions estimated via kernel smoothing and summarized through FPC analysis. |
| Thresholded network eigenvalues | Eigenvalues of thresholded $A$, $L$, $\hat{L}$ | Eigenvalues recomputed on networks pruned at an empirically optimized edge weight threshold, retaining only edges reflecting strong spatial proximity. |

All models except the demographics-only baseline included total time to completion, total ink time, total think time, and the image scaling factor as covariates. Time-based features are established predictors of cognitive outcomes in the dCDT literature and were included to ensure that network-based predictors were evaluated over and above these well-validated signals.^4,5^ The image scaling factor was included to account for variability in drawing size across participants that could otherwise confound the distance-based edge weights.

#### 2.6.2 Primary Analyses

All feature sets described in Table 1 (except for the graph convolutional network) were used as predictors in five-fold stratified cross-validation random forest models to predict cognitive status. Stratification ensured that the proportion of cognitively impaired participants was preserved across folds. Each model was evaluated separately under three conditions: command only, copy only, and combined command and copy, to assess whether predictive information varies by test conditions. Final model performance was assessed by aggregating predictions across the test folds.

Random forests were selected as the primary classification framework given their ability to handle high-dimensional, potentially correlated predictors, their robustness to distributional assumptions, and their established use in prior dCDT classification studies.^7,14^ To address the class imbalance in the sample (8.0% cognitively impaired), class-specific weights inversely proportional to empirical class frequencies were assigned during random forest training, upweighting observations in the minority (cognitively impaired) class relative to the majority (cognitively intact) class. Weights for each class *c* were calculated as *N_total_* /(*n_classes_* × *N_c_*).

Graph convolutional neural networks (GCNs) were also trained on the dCDT networks using twenty-three node-level features and a single edge feature representing inter-node spatial distances across three GCN layers. Node features included time to complete, bounding box height and width, bounding box area, convex hull area, number of strokes, mean and standard deviation of pressure, number of recorded points, drawing speed, and ink length, *x* and *y* coordinates of the node center, and a missingness indicator, as well as graph-level variables such as age, sex, APOE-*€*4 status, education, FHS cohort, image scale factor, total time to complete, total ink time, and total think time. The dataset was partitioned into the same five folds for cross-validation; in each fold, the model was trained on four folds using negative log-likelihood loss and validated on the remaining fold. Additional details on the GCN models are contained in the Supplementary Methods Section S1.1.

#### Secondary Analyses

Two additional exploratory approaches, edge thresholding and spectral density functions, were evaluated to examine whether alternative network representations could further improve classification performance. The edge thresholding approach assessed whether removing edges between distant nodes improves classification. An empirically optimized edge weight threshold was identified separately for command and copy networks. The spectral density functions approach computed network spectral density functions as a complementary representation of the eigenvalue distributions, summarized through functional principal component analysis.^25^ Full details for both approaches are provided in Supplementary Sections S1.2 and S1.3.

#### Performance Assessment

Model performance was assessed using AUC, sensitivity, and specificity. The decision threshold for sensitivity and specificity was determined using Youden’s index. All comparisons were performed within the same cross-validation folds. Confidence intervals for all performance metrics were estimated empirically across cross-validation folds. Statistical analyses were conducted in R using the packages `statGraph`, `caret`, `fdapace`, `qgraph`, and `igraph`, and in Python using `PyTorch`.

### 2.7 Subgraph Comparisons using Edit Distance

To identify localized structural differences between group-average networks, edit distances were computed for all possible four-node induced subgraphs. Four-node subgraphs were selected as the unit of comparison for two reasons. First, exhaustive enumeration over all 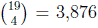 four-node induced subgraphs of the 19-node dCDT network is computationally tractable and provides dense spatial coverage. Second, four-node subgraphs are sufficiently small that each identified subgraph can be interpreted directly in terms of its constituent clock components, preserving the spatial interpretability that motivates the network representation. For each comparison, group-average networks were first constructed by computing mean edge weights across individuals within each cognitive group and test condition, and edit distances were then computed between the resulting average networks for all four-node induced subgraphs. Permutation testing was then conducted to assess the significance of differences in edit distances between all possible subgraphs. Details on the permutation tests are contained in the Supplementary Methods Section S1.4.

## 3 Results

### 3.1 Sample Characteristics

The final sample comprised 3,362 participants, of whom 3,092 (92.0%) were cognitively intact and 270 (8.0%) had cognitive impairment, including 193 with mild cognitive impairment and 77 with dementia of any cause (Supplementary Table S3). Cognitively impaired participants were older than intact participants (mean 81.1 [SD 7.9] vs. 62.6 [SD 13.3] years). Sex distribution was similar across groups (42.6% male among impaired vs. 45.9% among intact). College-level education was less prevalent among impaired participants (29.3%) than among intact participants (53.8%), and the APOE-ε4 allele was present in a higher proportion of the impaired group (28.9% vs. 22.3%). Cohort composition differed markedly between groups, reflecting the age distribution: the Offspring cohort represented 90.4% of impaired and 37.1% of intact participants, while the Third Generation cohort accounted for 53.0% of intact and 3.0% of impaired participants.

### 3.2 Prediction Model Performance

#### 3.2.1 Baseline Models

The demographics-only baseline achieved an AUC of 0.85 (95% CI: 0.84–0.87), with sensitivity of 0.85 (95% CI: 0.81–0.89) and specificity of 0.78 (95% CI: 0.76–0.79). Adding comprehensively collated existing summary features to demographic predictors slightly improved AUC to 0.86–0.87 across test conditions. Summary feature models were fitted on a slightly smaller sample (N = 3,267–3,343 depending on test condition) than the network-based models (N = 3,362), owing to missing values in some summary features when clock components were omitted from the drawing. Full performance metrics including AUC, sensitivity, and specificity scores are reported in Supplementary Table S4 and visualized in Figure 2.

**Figure 2:**
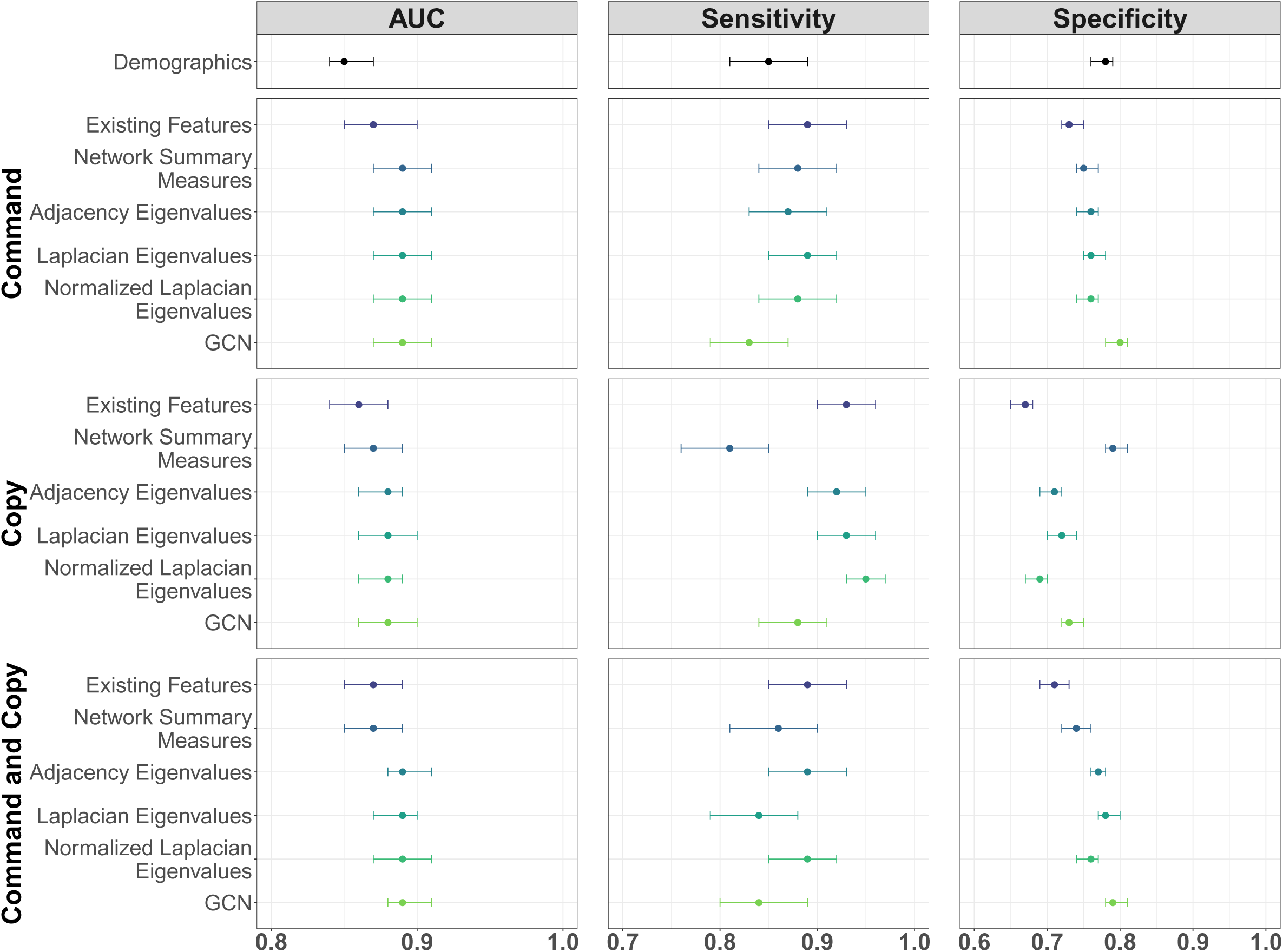
Cross-validated model performance for predicting cognitive impairment from dCDT-derived predictors. Points denote mean performance and error bars denote 95% confidence intervals estimated empirically across cross-validation folds. Metrics shown are area under the receiver operating characteristic curve (AUC), sensitivity, and specificity; sensitivity and specificity were computed using decision thresholds defined by Youden’s index. Results are shown separately for command-only, copy-only, and combined command and copy conditions. Full values are reported in Supplementary Table S4.

#### 3.2.2 dCDT Network-based Models

Network-based approaches, regardless of matrix representation or model, matched or slightly exceeded the comprehensively collated existing summary feature set across command, copy, and combined conditions, achieving AUCs of 0.87–0.89 with overlapping confidence intervals (Figure 2, Supplementary Table S4). In the command condition, network summary measures, all three eigenvalue-based representations, and the GCN achieved an AUC of 0.89 (95% CI: 0.87–0.91). In the copy condition, all network models achieved AUC values of 0.87–0.88, slightly higher than the summary feature baseline of 0.86 (95% CI: 0.84–0.88). In the combined command and copy condition, all three eigenvalue-based representations and the GCN yielded the highest AUC at 0.89 (95% CI: 0.88–0.91).

At the Youden’s index threshold, a sensitivity-specificity trade-off was generally observed across network representations and test conditions. In the command condition, the eigenvalue models had specificity near 0.76 and sensitivity between 0.87 and 0.89, whereas the GCN showed lower sensitivity of 0.83 (95% CI: 0.79–0.87) but higher specificity of 0.80 (95% CI: 0.78–0.81). In the copy condition, the eigenvalue models achieved the highest sensitivity, whereas the GCN showed lower sensitivity and similar specificity (0.73 vs. 0.69–0.72). In the combined condition, the GCN and Laplacian models achieved lower sensitivities and higher specificities than the adjacency matrix and normalized Laplacian models.

#### 3.2.3 Secondary Analyses

Spectral density functions of the adjacency, Laplacian, and normalized Laplacian matrices, summarized via functional principal component analysis, yielded AUC values of 0.86–0.89 across test conditions (Supplementary Table S5), similar to or marginally lower than the corresponding unthresholded eigenvalue models with no consistent improvement in sensitivity or specificity.

For network thresholding, optimal edge weight thresholds of 0.70 and 0.80 were identified for command and copy networks, respectively, corresponding to the removal of edges with standardized Euclidean distances greater than 0.36 and 0.22. The thresholded networks yielded slightly lower AUC values of 0.84–0.88 across all three test conditions and matrix representations (Supplementary Table S6) compared to the unthresholded networks.

Confidence intervals for thresholded models were wider than for their unthresholded counterparts. The distribution of optimal threshold values across 10,000 training set splits is displayed in Supplementary Figure S1.

### 3.3 Network Structure and Cognitive Status Differences

Group-level average networks by cognitive status and test condition are shown in Figure 3. Structural summaries of the networks by group and condition are reported in Supplementary Table S7. In the command condition, 28.1% of cognitively impaired participants had at least one isolated node compared with 4.7% of intact participants, and mean node degree variability was higher in the impaired group (1.21 [SD 1.14] vs. 0.64 [SD 0.47]). Both differences were attenuated in the copy condition, where isolated nodes were present in 9.6% of impaired and 1.1% of intact participants.

**Figure 3:**
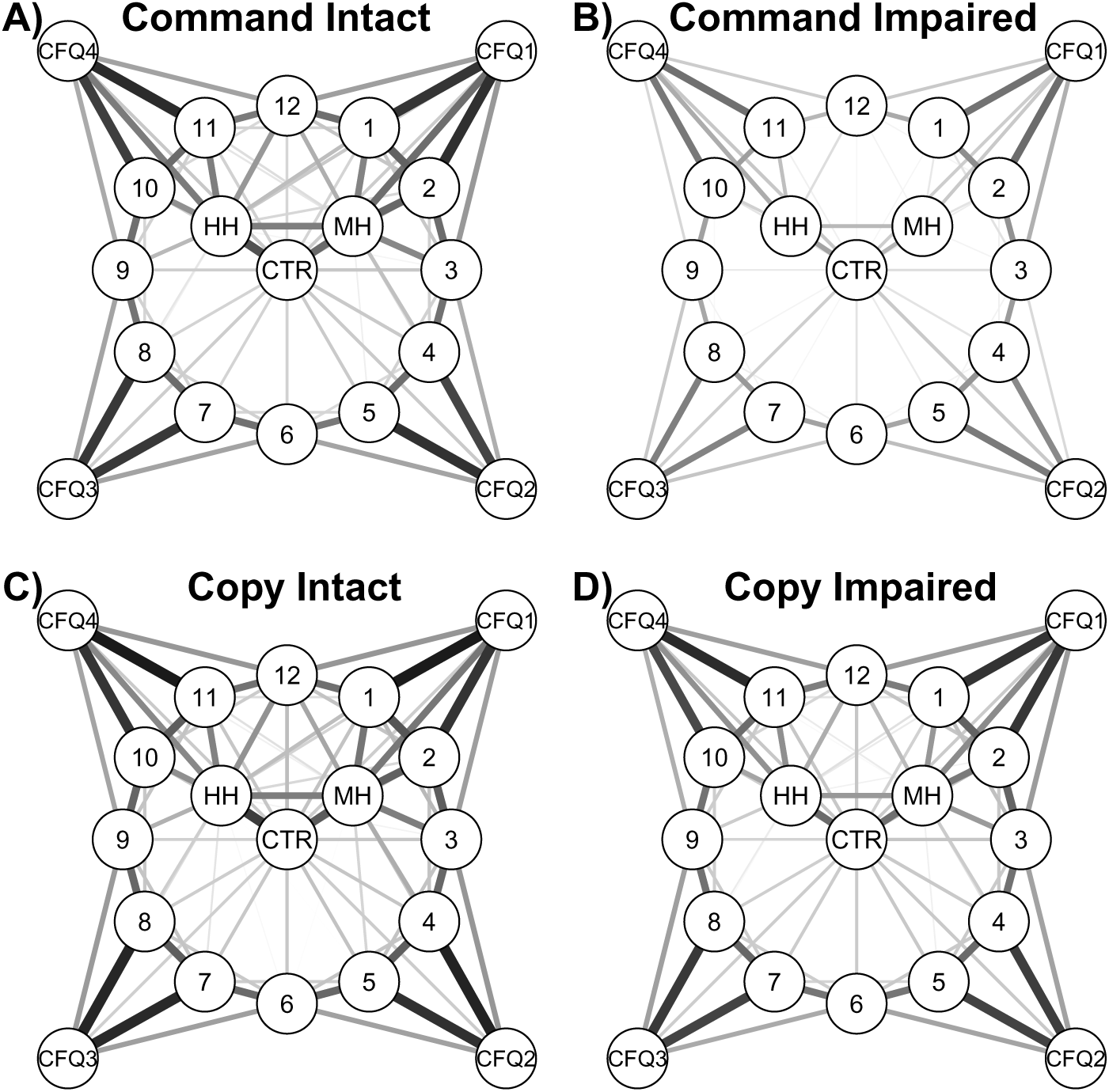
Group-average dCDT network representations by cognitive status and test condition. Panels show A) command intact, B) command impaired (top row), C) copy intact, and D) copy impaired (bottom row). Average networks were constructed by computing the mean edge weight across participants within each cognitive group. Line thickness is proportional to mean edge weight, with thicker edges indicating greater spatial proximity between components. Edges with mean weight below 0.60 (corresponding to mean standardized Euclidean distance greater than 0.51) are removed for visualization. Node labels: HH = Hour Hand; MH = Minute Hand; CFQ1–CFQ4 = Clock Face Quadrants 1–4; CTR = Clock Face Center.

#### 3.3.1 Comparisons Between Cognitive Status Groups

Figure 4 contains the top three subgraphs with the largest edit distances between cognitive groups in the command and copy conditions. In the command condition, the three subgraphs with the largest between-group edit distances were *V*_1_^Comm^ = {1, 2, MH, CF Q1}, *V*_2_^Comm^ = {1, 2, HH, MH}, and *V*_3_^Comm^ = {2, HH, MH, CF Q1}, with edit distances of 0.67, 0.66, and 0.64, respectively (*p* < 0.001). The largest differences involved components located in the upper right-hand region of the clock: digits 1 and 2, the clock hands, and the first clock face quadrant. In the copy condition, the three subgraphs with the largest between-group edit distances were *V*_1_^Copy^ = {1, HH, MH, CF Center}, *V*_2_^Copy^ = {4, MH, HH, CF Q2}, and *V*_3_^Copy^ = {1, 4, MH, HH}, with edit distances of 0.25, 0.24, and 0.24, respectively (*p* < 0.001). Between-group edit distances in the copy condition were considerably smaller than in the command condition, and the identified subgraphs extended toward digit 4 and the second clock face quadrant.

**Figure 4:**
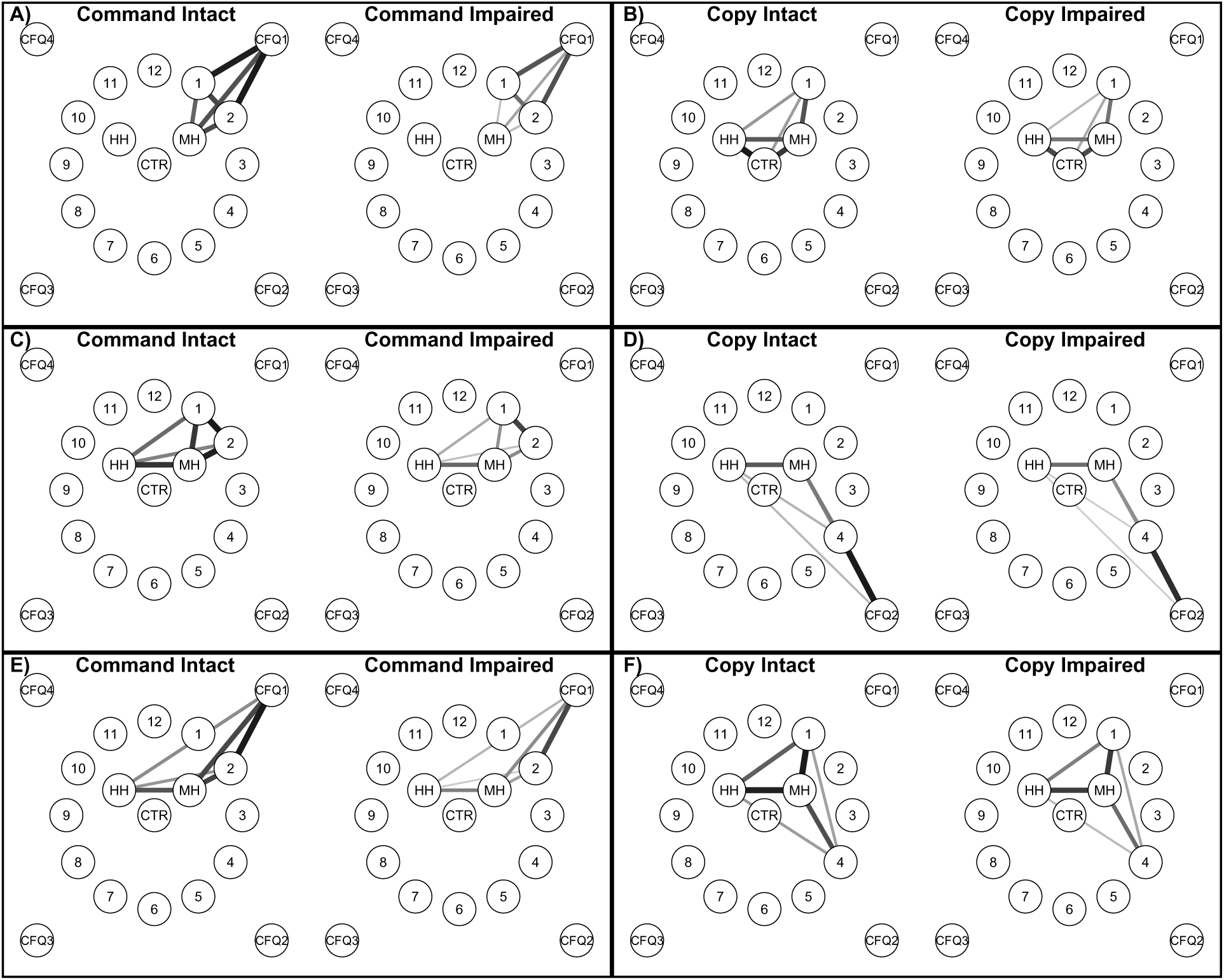
Localized between-group network differences identified by four-node subgraph edit distance. Cognitively intact and impaired group-average networks were compared by computing edit distances for all possible four-node induced subgraphs. The three subgraphs with the largest edit distances are displayed for the command (A, C, E) and copy (B, D, F) conditions, highlighting localized component configurations that differ most between cognitive groups. Node labels: HH = Hour Hand; MH = Minute Hand; CFQ1–CFQ4 = Clock Face Quadrants 1–4; CTR = Clock Face Center.

#### 3.3.2 Within-Group Comparisons Between Test Conditions

Figure 5 contains the top three subgraphs with the largest edit distances between test conditions within each cognitive group. In the cognitively intact group, the three subgraphs with the largest command-to-copy edit distances were *V*_1_^Intact^ = {1, 4, MH, CF Q2}, *V*_2_^Intact^ = {4, 6, MH, CF Q2}, and *V*_3_^Intact^ = {4, 5, 6, MH}, with edit distances of 0.18, 0.18, and 0.17, respectively (*p* < 0.001). In the cognitively impaired group, the three subgraphs with the largest command-to-copy edit distances were *V*_1_^Impaired^ = {2, 4, MH, CF Q2}, *V*_2_^Impaired^ = {2, 3, MH, CF Q2}, and *V*_3_^Impaired^ = {3, 4, MH, CF Q2}, with edit distances of 0.62, 0.60, and 0.60, respectively (*p* < 0.001). Command-to-copy edit distances in the impaired group were approximately 3.4-fold larger than in the intact group; the most-differentiating subgraphs in the impaired group involved digits 2 through 4, the minute hand, and clock face quadrant 2.

**Figure 5:**
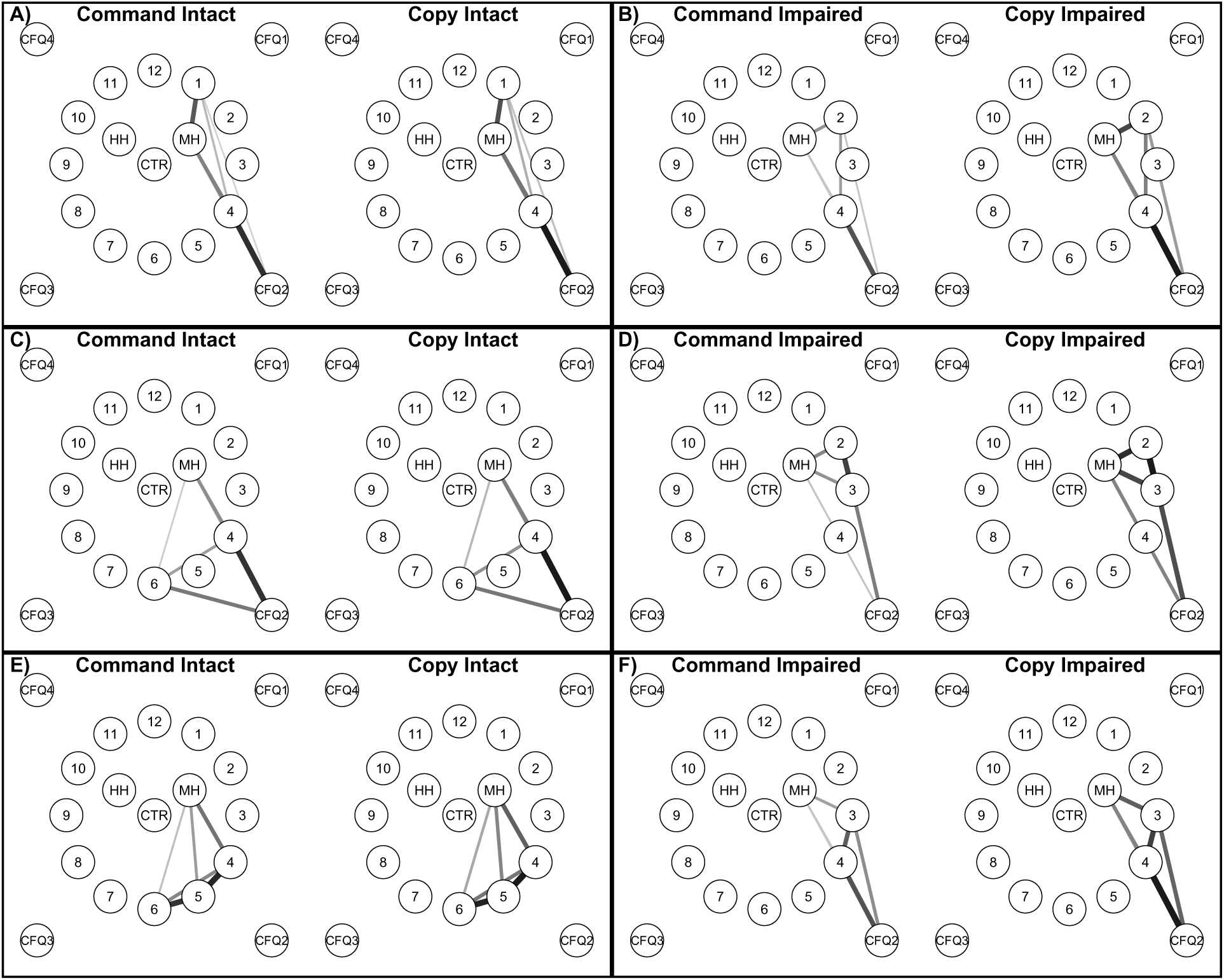
Localized within-group command–copy differences identified by four-node subgraph edit distance. Command and copy average networks were compared by computing edit distances for all possible four-node induced subgraphs. The three subgraphs with the largest edit distances are displayed for the cognitively intact group (A, C, E) and impaired group (B, D, F), highlighting localized component configurations that differ most between between test conditions. Node labels: HH = Hour Hand; MH = Minute Hand; CFQ1– CFQ4 = Clock Face Quadrants 1–4; CTR = Clock Face Center.

## 4 Discussion

dCDT networks encode the organization of the drawing by capturing spatial relationships among clock components, including omitted or misplaced clock components. Our study demonstrates that structural network representations of the dCDT achieve predictive performance comparable to or slightly better than established summary features for classifying cognitive impairment, while offering several interpretive and methodological advantages. This finding is noteworthy because the summary features used for comparison are a comprehensive collation of existing features in the literature designed to capture drawing efficiency, motor coordination, and spatial accuracy. The fact that our fully automated, component-level spatial representation matches this performance without any investigator-driven feature selection supports the practical value and scalability of the proposed approach.

The demographics-only model provided a strong predictive baseline (AUC 0.85). Adding the comprehensive set of summary features resulted in only marginal improvement (AUC 0.86–0.87), and the network-based models yielded modest further gains (AUC 0.87–0.89). In this sample, demographics captured much of the predictive signal, likely because cognitively impaired participants were on average about 18 years older than intact participants and were mostly from the Offspring cohort. This leaves little room for any drawing-based measure to improve the AUC. Thus, the AUC may underestimate the added value of dCDT features, particularly if they capture patterns not explained by demographics. It also means that the size of these improvements may be specific to the FHS sample. Replication in cohorts with different age, education, and demographic profiles could provide further insights.

### 4.1 Comparison Across Network Summaries

Among the different matrix representations of the networks evaluated, no single representation consistently outperformed the others in terms of overall AUC. This suggests that the structural information encoded by the eigenvalue-based representations and the GCN is largely overlapping in this context. Observed differences were primarily in the sensitivity and specificity values. Under Youden’s index, the eigenvalue-based models generally yielded higher sensitivity and lower specificity, whereas the GCN yielded higher specificity and lower sensitivity. These differences were modest, but they suggest that the choice of model may be guided by whether the clinical objective prioritizes sensitivity or specificity.

Spectral density functions and eigenvalues from the thresholded networks did not offer meaningful improvements over eigenvalues from the unthresholded networks in the classification models evaluated here. The lack of advantage from spectral densities may reflect the spatially derived and weighted nature of the dCDT networks, which differs structurally from the unweighted, randomly generated networks for which spectral density discrimination methods were originally developed.^26^ Similarly, edge thresholding did not consistently improve, and in some cases marginally reduced, model performance, possibly because low-weight edges reflecting distant clock components carry information about spatial disorganization that is itself predictive of cognitive impairment. Additionally, thresholding does not differentiate between low-weight edges and absent edges, potentially discarding relevant information. The larger confidence interval lengths for the thresholded models also indicate greater uncertainty, likely attributable to the small held-out validation set used for threshold selection.

### 4.2 Differences Across Test Conditions and Cognitive Groups

Beyond classification performance, the network representation enables direct localization of structural differences between cognitive groups in a way that scalar summary features cannot. In the command condition, the three four-node subgraphs with the largest edit distances between the group-average networks (0.64 to 0.67) consistently involved digits 1 and 2, the clock hands, and the first quadrant of the clock face (Figure 4). These are precisely the components required to accurately represent the target time used in this study. The finding that these components are, on average, spaced farther apart in the cognitively impaired group suggests that impaired participants have greater difficulty positioning the upper-right numerals and clock hands in their correct spatial relationships, consistent with known deficits in visuospatial processing and executive function associated with cognitive decline. This degree of spatial specificity, identifying not just that the drawing differs between groups but where it differs, is not accessible from summary statistics alone.

The copy condition exhibited notably lower edit distances in the three most distinct subgraphs (0.24 to 0.25), with the identified subgraphs extending from digit 1 to the lower-right region of the clock face (Figure 4). This reduction in between-group structural differences when a visual reference is provided suggests that the spatial disorganization observed in the command condition reflects impairment in internally guided spatial reconstruction rather than fundamental deficits in motor execution. This distinction between internally guided and externally supported performance is consistent with the role of executive function and working memory in visuospatial planning.

Examining within-group differences between test conditions revealed a clinically meaningful asymmetry between cognitive groups (Figure 5). Cognitively intact participants maintained highly consistent spatial organization across test conditions, with the three largest edit distances between their average command and copy networks ranging from only 0.17 to 0.18. In contrast, cognitively impaired participants exhibited substantially larger structural shifts between conditions, with edit distances of 0.60 to 0.62 in the three most differentiating subgraphs. Specifically, the mid-to-lower right region of the clock, involving digits 2 through 4, the minute hand, and clock face quadrant 2, was drawn with greater spatial spread in the command condition than in the copy condition among impaired participants. This pattern suggests that a visual reference substantially reorganizes the spatial output of cognitively impaired individuals in a way that does not occur in intact participants, consistent with the well-documented benefit of external cueing on task performance in individuals with executive dysfunction.^27^ The network representation makes this spatial reorganization directly quantifiable and regionally localizable, providing a level of interpretive resolution that is not achievable through aggregate summary measures.

### 4.3 Strengths, Limitations, and Future Directions

The proposed network representation offers several practical advantages over summary feature-based approaches. First, it is fully objective; once the node set and edge weighting function are defined, no further subjective decisions are required, removing the feature selection and correlation management steps. Second, the representation handles missing clock components naturally, as an omitted component results in an isolated node rather than a missing value, preserving the structural information that the component was absent without requiring imputation or exclusion. Third, the full spatial geometry of the drawing is retained at the component level, providing a richer computational object than a fixed-length feature vector. Finally, the representation is directly extensible as (a) directed edges could encode the temporal sequence of component drawing, (b) edge weights could reflect drawing duration rather than spatial distance, and (c) the node set could be expanded to capture finer spatial subdivisions of the clock face. Such extensions are not feasible within the scalar summary feature framework.

Some limitations of this study warrant acknowledgment. The FHS sample is predominantly White and highly educated, which limits the generalizability of these findings. Additionally, all performance estimates are derived from cross-validation within a single cohort; the absence of external validation further limits generalizability. The cross-sectional design, using only the most recent dCDT per participant, precludes conclusions about whether network features predict future cognitive decline in currently intact individuals, which represents an important and directly testable extension of this work. Cognitively intact status was assumed for participants who had not been flagged through the dementia review process; because cognitive impairment can go undetected until years after symptom onset, some misclassification of intact participants is possible. Finally, the subgraph comparisons were based on group-average networks. Although permutation tests with correction for multiple comparisons indicated that the largest differences were unlikely to be due to chance, these comparisons only describe average patterns and do not show how well each subgraph discriminates between individual participants.

Future directions include longitudinal analysis examining whether baseline network features are associated with subsequent cognitive decline in participants who are cognitively intact at the time of their dCDT, which would directly address the clinical utility of the proposed representation for early detection. Methods for testing differences in whole-network topology, and for linking subgraph differences to individual-level prediction, would further strengthen the inference. Extensions to directed and temporally weighted networks could capture sequential and timing aspects of the drawing process not encoded in the current spatial representation.

The dCDT network representation offers potential to enhance clinical assessment by providing an objective, interpretable, and scalable complement to standard dCDT evaluation. After external validation, this approach could be incorporated into a clinical workflow in which patients complete the dCDT and the drawing is converted automatically into a network representation. Specialized software would then return a predicted risk of cognitive impairment together with a visual summary of the clock regions that differ most from typical drawings.

The sensitivity-specificity trade-off observed across models also suggests potential utility for different clinical applications. Eigenvalue-based models, which are computationally efficient for small networks and demonstrated higher sensitivity, may be useful in initial screening settings where minimizing missed cases of cognitive impairment is the priority. In contrast, GCN models, which are more computationally intensive and demonstrated higher specificity, may be more appropriate for follow-up evaluations, where fewer individuals require assessment and reducing false positives is the priority.

## Conflicts

Rhoda Au reports serving as a consultant to Signant Health.

## Funding

This work was supported by the Framingham Heart Study’s National Heart, Lung, and Blood Institute contract (N01-HC-25195), and by grants from the National Institute of Neurological Disorders and Stroke (R01-NS017950), the National Institute on Aging (U19-AG068753; 1R03AG098551-01; AG062109; AG072654; AG083735) and the Framingham Heart Study Brain Aging Program Pilot Award (U19-AG068753).

## Consent statement

The Institutional Review Board of Boston University Medical Center approved the study protocol and informed consent was obtained from all participants.

## Data Availability

Framingham Heart Study data are available to qualified researchers through application to the Framingham Heart Study [https://www.framinghamheartstudy.org/fhs-for-researchers/] and via NHLBI's BioLINCC and dbGaP repositories, subject to data use agreements. Digital clock drawing data are available through the Framingham Heart Study research application process. The code used to convert digital pen data to structural networks is publicly available at https://github.com/shariq-mohammed/clocknet.

## Notes

### Competing Interest Statement

Rhoda Au reports serving as a consultant to Signant Health. All other authors declare no competing interests, and none of the authors or their institutions received payments or services from a third party in the past 36 months that could be perceived to influence the submitted work.

