## Supplementary Materials for "Network representations of digital clock drawings for cognitive impairment detection"

### S1 Supplementary Methods

#### S1.1 Graph Convolutional Network

The command and copy networks were integrated into a single model as a single graph formed by the disjoint union of the command and copy networks. Command and copy nodes form two separate, non-overlapping node sets, with edges connecting nodes within each set and no edges linking the command and copy node sets.

The GCN model consisted of three graph convolutional layers, each with 64 hidden channels. The first graph convolutional layer used ReLU activation and dropout probability of 0.20, while the subsequent layers used ReLU activation without dropout. Node and edge information were incorporated into the node representations and a residual connection was used to combine edge-informed features with the graph-convolutional node embeddings. The resulting node embeddings were summarized into a graph-level representation using global mean pooling. This graph representation was passed through a fully connected layer with ReLU activation with dropout probability of 0.50 and batch normalization, followed by a final linear output layer.

Models were trained and evaluated using 5-fold cross-validation. For each fold, models were optimized using an Adam optimizer with a learning rate of 0.01 and weight decay of $5 \times 10^{-4}$ for $L_2$ regularization. All models were trained for a fixed 500 epochs on the training data. Final models were evaluated on held-out test sets, and results were aggregated across all five folds to produce pooled out-of-fold performance estimates. Hyperparameters were determined based on prior work [1, 2].

#### S1.2 Edge-Thresholding

Edge-thresholding was evaluated as an exploratory approach to assess whether removing weak (low-weight) edges between spatially distant clock components improves classification performance. Networks were randomly divided into a training set (80%) and a held-out validation set (20%). Within the training set, a candidate edge-weight threshold was applied to prune edges below the threshold, and eigenvalues were recomputed for the resulting adjacency, Laplacian, and normalized Laplacian matrices. The recomputed eigenvalues, together with covariates, were used to train random forest classifiers using five-fold cross-validation. Model discrimination was quantified using area under the receiver operating characteristic curve (AUC).

This threshold-selection procedure was repeated across a grid of candidate thresholds to identify the threshold maximizing cross-validated AUC. To characterize variability in the selected threshold, the process was repeated across 10,000 resampled five-fold splits of the training data, yielding a distribution of optimal thresholds; the mode of this distribution was selected as the final threshold for evaluation in the held-out validation set.

#### S1.3 Spectral Density Functions

The *spectral density* of a graph matrix is the distribution of the eigenvalues obtained from its eigendecomposition. For a graph $G$ with $N$ nodes and corresponding eigenvalues $\lambda_1 \geq \lambda_2 \geq ... \geq \lambda_N$, the spectral density is defined as $\rho(\lambda) = \frac{1}{N}\sum_{j=1}^{N} \delta(\lambda - \lambda_j)$, where $\delta$ is the Dirac delta function. The spectral density of the adjacency matrix can be directly related to the graph's topological features [3]. The empirical density can be estimated using established methods by applying Gaussian kernel regression with the Nadaraya-Watson estimator for smoothing, followed by normalization to ensure the area under the curve equals one.[4]

Network spectral density functions were incorporated into the models by applying functional principal components analysis (FPCA) via the Principal Analysis by Conditional Expectation (PACE) algorithm to capture the underlying structure of the functions. [5] PACE is well-suited for longitudinal data with irregular and sparse observations, as it estimates the mean and covariance structure of the functional data without requiring pre-smoothing of the individual functions. It reconstructs individual trajectories and extracts principal components that represent dominant modes of variation across participants.

Within each cross-validation fold, PACE was applied to the spectral density functions from the training set to estimate the eigenfunctions from the FPCA. The spectral density functions from the validation set were then projected onto these training-derived eigenfunctions to obtain principal component scores. The principal component scores were then used as input features along with participant demographics and image scaling factor in random forest models to predict cognitive status. The number of components retained in the classification models was selected to account for at least 99% of the total variability in the spectral density functions.

#### S1.4 Subgraph Permutation Testing

A permutation-based subgraph analysis was used to assess group differences in local network structure between cognitive groups and between test conditions. For a fixed subgraph size of four nodes, all possible four-node combinations from the 19-node network were enumerated.

Within each permutation, group labels were randomly shuffled without replacement, preserving the original group sizes. Group-specific mean adjacency matrices were then computed by averaging the weighted adjacency matrices within each permuted group. For each induced four-node subgraph, the edit distance between the corresponding group-level adjacency matrices was calculated. This procedure was repeated for 10,000 permutations to generate an empirical null distribution of subgraph-level group differences. Empirical p-values were then adjusted for multiple comparisons using the Benjamini–Hochberg procedure.

To assess differences between test conditions, condition labels were randomly flipped within each participant with probability 0.5 at each permutation, preserving the paired structure of the data. The permutation-based subgraph analysis then proceeded as described above.

### S2 Supplementary Tables

**Table S1:** Definitions of the 47 dCDT-derived summary features constituting the comprehensive summary-feature benchmark used in prediction models. Features are organized by domain: drawing efficiency, information processing, simple and complex motor skills, and spatial reasoning. Source citations correspond to published studies from which each feature was identified.

| Feature | Description | Source |
|---|---|---|
| ***Drawing efficiency*** | | |
| Number of strokes | Total number of strokes used to complete the test | [6, 7] |
| Total time | Total time to complete the test | [6, 8–10] |
| Ink length | Sum of the length of all strokes in the test | [6–8] |
| Strokes per minute | Total number of strokes divided by time | [9] |
| Clock face number of strokes | Total number of strokes used to complete the clock face | [6, 7] |
| Clock face total time | Total time to complete the clock face | [6] |
| Velocity | Drawing ink length divided by total time | [6, 9] |
| Stroke count conformity | The deviation from the expected number of pen strokes in the drawing | [8] |
| ***Information processing*** | | |
| Think time | The amount of time not spent drawing, calculated by subtracting the ink time from total time | [9] |
| Percent think time | Think time divided by total time | [8] |
| Average latency | The average duration of time between each pen stroke | [8] |
| Latency variability | Standard deviation of time between each pen stroke | [8] |
| Post-clock face latency | Delay between finishing the last stroke in the clock face and whatever is drawn next (sec) | [6, 10, 11] |
| Pre-first hand latency | Delay between whatever was drawn before the first clock hand and starting to draw that clock hand (sec) | [6, 10–12] |
| Pre-second hand latency | Delay between whatever was drawn before the second clock hand and starting to draw that clock hand (sec) | [6, 11] |
| Longest latency | The longest time between the end of one stroke and whatever was drawn next | [8] |
| Relative long latency | A measure of the difference between the average latency and the longer latencies within this drawing | [8] |

| Feature | Description | Source |
|---|---|---|
| Long latency count | The total number of latencies in the drawing that are notably longer than the normative sample standard. | [8] |
| ***Simple and complex motor*** | | |
| Ink time | Total time with the pen on the paper | [6, 9] |
| Percent ink time | Total time with the pen on the paper divided by total time to complete the test | [8] |
| Clock face average speed | Average speed of the pen for all strokes used to complete the clock face | [8] |
| Clock face maximum speed | Maximum speed of the pen when drawing the clock face | [8] |
| Clock face initiation speed | Speed of the pen at the beginning of drawing the clock face | [8] |
| Clock face termination speed | Speed of the pen at the end of drawing the clock face | [8] |
| Average pen pressure | Average pen pressure of all the points in the test | [9] |
| Standard deviation of pressure | Standard deviation of pressure for all points in the test | [6] |
| Average pressure/velocity | Ratio of average pressure to average velocity | [6, 9] |
| ***Spatial reasoning*** | | |
| Missing digits | Any digits not appearing on the test | [6] |
| Digit perseveration | Any repeated digits | [6] |
| Missing hands | Missing hour or minute hand | [6] |
| Digit over 12 | Any digits greater than 12 | [6] |
| Hand perseveration | Any repeated minute or hour hand | [6] |
| Hand crossed out | Any hour hand or minute hand that is crossed out | [6] |
| Anchoring | Starting with digits 3,6,9,12 (in any order) before drawing all other digits | [6, 13] |
| Ten-eleven stroke | A line connecting digits 10 and 11 | [6] |
| Clock face area | Area of a circle ($\pi r^2$) using the average radius of horizontal and vertical radii | [6, 10] |
| Hour hand length | Ink length of the hour hand | [6] |
| Hour hand distance from centroid | Distance between the closest point on the hour hand to the centroid of the clock face | [6] |
| Minute hand length | Ink length of the minute hand | [6] |
| Minute hand distance from centroid | Distance between the closest point on the minute hand to the centroid of the clock face | [6] |
| Ratio of minute to hour hand length | Length of minute hand divided by length of hour hand | [6] |
| Average digit height | The average of the heights of all digits drawn on the test | [6] |

| **Feature** | **Description** | **Source** |
|---|---|---|
| Average digit width | The average of the widths of all digits drawn on the test | [6] |
| Average digit distance from circumference | The average distance from the closest point of each digit to the clock face | [6] |
| Clock face overshoot distance | The minimum distance between the start and end of the clock face divided by the total length of the clock face | [6] |
| Symmetry | Ratio of the length to width of the clock face | [6, 8] |
| Correctly placed hands | Minute and hour hand pointing to the correct digits | [14] |

**Table S2:** Definitions of graph-theoretic network summary measures used as predictors in random forest classification models. Measures capture global and local structural properties of participant-level dCDT networks, including degree-based summaries, density, centrality, distance, and clique-based statistics.

| **Feature** | **Description** |
|---|---|
| Number of edges | Total number of edges connecting all nodes |
| Edge density | Ratio of the total number of edges and the largest number of possible edges |
| Number of isolated nodes | Number of nodes with no edge |
| Mean edge weight | Average of all edge weights |
| Minimum edge weight | Minimum value of all edge weights |
| Maximum edge weight | Maximum value of all edge weights |
| Mean node degree | Average number of edges across nodes |
| Mean weighted node degree | Average sum of the weights of all edges connected to each node |
| Node degree variability | Standard deviation of node degrees across all nodes |
| Weighted node degree variability | Standard deviation of the sum of the edge weights for all nodes |
| Maximum node degree | The largest number of connections a node has across all nodes |
| Minimum node degree | The smallest number of connections a node has across all nodes |
| Maximum weighted degree | The largest sum of weights of all edges for a node across all nodes |
| Minimum weighted degree | The smallest sum of weights of all edges for a node across all nodes |
| Diameter | The largest shortest path length (sum of the edge weights) between any two nodes |
| Mean distance | The average of the shortest path lengths between all pairs of nodes in the network |
| Size of the largest clique | The number of nodes in the largest possible clique (a subgraph where every node is connected to all other nodes) |
| Count of maximal cliques | The number of cliques that are not contained in any larger clique |
| Mean centrality | Average power centrality (node influence) across all nodes |
| Mean eccentricity | Average eccentricity (shortest path distance from the farthest other node in the graph) across all nodes |
| Radius | The smallest eccentricity across all nodes |
| Mean harmonic centrality | Average harmonic centrality (mean inverse distance to all other vertices) across all nodes |

**Table S3:** Participant characteristics overall and stratified by cognitive status. Values are mean (SD) for continuous variables and $n$ (%) for categorical variables; percentages are column percentages. APOE-$\varepsilon$4+ indicates presence of at least one $\varepsilon$4 allele. Cognitive impairment is defined as MCI or all-cause dementia.

| | **Overall** ($N = 3,362$) | **Cognitively Intact** ($N = 3,092$) | **Dementia or MCI** ($N = 270$) |
|---|---|---|---|
| Age, mean (SD) | 64.1 (13.9) | 62.6 (13.3) | 81.1 (7.86) |
| Male, n (%) | 1535 (45.7%) | 1420 (45.9%) | 115 (42.6%) |
| Education, n (%) | | | |
| Less than high school | 63 (1.9%) | 48 (1.6%) | 15 (5.6%) |
| High school | 531 (15.8%) | 454 (14.7%) | 77 (28.5%) |
| Some college | 1027 (30.5%) | 928 (30.0%) | 99 (36.7%) |
| College or more | 1741 (51.8%) | 1662 (53.8%) | 79 (29.3%) |
| Framingham cohort, n (%) | | | |
| Original | 9 (0.3%) | 3 (0.1%) | 6 (2.2%) |
| Offspring | 1391 (41.4%) | 1147 (37.1%) | 244 (90.4%) |
| New Offspring Spouse | 33 (1.0%) | 32 (1.0%) | 1 (0.4%) |
| Third Generation | 1648 (49.0%) | 1640 (53.0%) | 8 (3.0%) |
| Omni 1 | 163 (4.8%) | 153 (4.9%) | 10 (3.7%) |
| Omni 2 | 118 (3.5%) | 117 (3.8%) | 1 (0.4%) |
| APOE4+, n (%) | 766 (22.8%) | 688 (22.3%) | 78 (28.9%) |
| All-cause dementia, n(%) | - | - | 77 (28.5%) |
| Mild cognitive impairment, n(%) | - | - | 193 (71.5%) |

**Table S4:** Cross-validated performance of primary dCDT classification models for predicting cognitive impairment (mild cognitive impairment or all-cause dementia). Sensitivity and specificity were computed using decision thresholds defined by Youden's index. Sample size ($n$) reflects participants with complete data for the corresponding feature set and condition. All models include demographic covariates (age, sex, education, Framingham cohort, and APOE-$\varepsilon$4 carrier status); all models except the demographics-only baseline additionally include total time to completion, total ink time, total think time, and image scaling factor.

| **Model** | **n** | **AUC (95% CI)** | **Sensitivity (95% CI)** | **Specificity (95% CI)** |
|---|---|---|---|---|
| Demographics | 3362 | 0.85 (0.84-0.87) | 0.85 (0.81-0.89) | 0.78 (0.76-0.79) |
| *Command Only* | | | | |
| Existing summary features | 3273 | 0.87 (0.85-0.90) | 0.89 (0.85-0.93) | 0.73 (0.72-0.75) |
| Network summary measures | 3362 | 0.89 (0.87-0.91) | 0.88 (0.84-0.92) | 0.75 (0.74-0.77) |
| Adjacency Eigenvalues | 3362 | 0.89 (0.87-0.91) | 0.87 (0.83-0.91) | 0.76 (0.74-0.77) |
| Laplacian Eigenvalues | 3362 | 0.89 (0.87-0.91) | 0.89 (0.85-0.92) | 0.76 (0.75-0.78) |
| Normalized Laplacian Eigenvalues | 3362 | 0.89 (0.87-0.91) | 0.88 (0.84-0.92) | 0.76 (0.74-0.77) |
| GCN | 3362 | 0.89 (0.87-0.91) | 0.83 (0.79-0.87) | 0.80 (0.78-0.81) |
| *Copy Only* | | | | |
| Existing summary features | 3343 | 0.86 (0.84-0.88) | 0.93 (0.90-0.96) | 0.67 (0.65-0.68) |
| Network summary measures | 3362 | 0.87 (0.85-0.89) | 0.81 (0.76-0.85) | 0.79 (0.78-0.81) |
| Adjacency Eigenvalues | 3362 | 0.88 (0.86-0.89) | 0.92 (0.89-0.95) | 0.71 (0.69-0.72) |
| Laplacian Eigenvalues | 3362 | 0.88 (0.86-0.90) | 0.93 (0.90-0.96) | 0.72 (0.70-0.74) |
| Normalized Laplacian Eigenvalues | 3362 | 0.88 (0.86-0.89) | 0.95 (0.93-0.97) | 0.69 (0.67-0.70) |
| GCN | 3362 | 0.88 (0.86-0.90) | 0.88 (0.84-0.91) | 0.73 (0.72-0.75) |
| *Command and Copy* | | | | |
| Existing summary features | 3267 | 0.87 (0.85-0.89) | 0.89 (0.85-0.93) | 0.71 (0.69-0.73) |
| Network summary measures | 3362 | 0.87 (0.85-0.89) | 0.86 (0.81-0.90) | 0.74 (0.72-0.76) |
| Adjacency Eigenvalues | 3362 | 0.89 (0.88-0.91) | 0.89 (0.85-0.93) | 0.77 (0.76-0.78) |
| Laplacian Eigenvalues | 3362 | 0.89 (0.87-0.90) | 0.84 (0.79-0.88) | 0.78 (0.77-0.80) |
| Normalized Laplacian Eigenvalues | 3362 | 0.89 (0.87-0.91) | 0.89 (0.85-0.92) | 0.76 (0.74-0.77) |
| GCN | 3362 | 0.89 (0.88-0.91) | 0.84 (0.80-0.89) | 0.79 (0.78-0.81) |

**Table S5:** Cross-validated performance of dCDT classification models using spectral density function representations (secondary analysis). Spectral density functions of network matrix eigenvalues were estimated via Gaussian kernel smoothing and summarized through FPCA; retained components accounted for at least 99% of total variability. Sensitivity and specificity were computed using Youden's index. All models include the same covariates as the primary network-based models (Supplementary Table S4).

| **Model** | **n** | **AUC (95% CI)** | **Sensitivity (95% CI)** | **Specificity (95% CI)** |
|---|---|---|---|---|
| *Command Only* | | | | |
| Adjacency Spectral Density | 3362 | 0.88 (0.86-0.90) | 0.92 (0.89-0.95) | 0.69 (0.68-0.71) |
| Laplacian Spectral Density | 3362 | 0.88 (0.86-0.90) | 0.92 (0.89-0.95) | 0.71 (0.69-0.73) |
| Normalized Laplacian Spectral Density | 3362 | 0.88 (0.86-0.90) | 0.85 (0.81-0.89) | 0.78 (0.77-0.80) |
| *Copy Only* | | | | |
| Adjacency Spectral Density | 3386 | 0.87 (0.85-0.89) | 0.81 (0.77-0.86) | 0.77 (0.75-0.78) |
| Laplacian Spectral Density | 3386 | 0.88 (0.86-0.90) | 0.87 (0.82-0.91) | 0.74 (0.73-0.76) |
| Normalized Laplacian Spectral Density | 3386 | 0.86 (0.84-0.88) | 0.88 (0.84-0.91) | 0.71 (0.69-0.72) |
| *Command and Copy* | | | | |
| Adjacency Spectral Density | 3362 | 0.89 (0.87-0.91) | 0.83 (0.78-0.87) | 0.80 (0.78-0.81) |
| Laplacian Spectral Density | 3362 | 0.88 (0.86-0.90) | 0.89 (0.85-0.93) | 0.75 (0.73-0.76) |
| Normalized Laplacian Spectral Density | 3362 | 0.88 (0.86-0.90) | 0.80 (0.75-0.85) | 0.80 (0.78-0.81) |

**Table S6:** Cross-validated performance of dCDT classification models using edge-thresholded network representations (secondary analysis). Eigenvalues were recomputed on networks pruned at empirically optimized edge-weight cutoffs: 0.70 for command networks and 0.80 for copy networks, corresponding to removal of edges with standardized Euclidean distances greater than 0.36 and 0.22, respectively. All models include the same covariates as the primary network-based models (Supplementary Table S4).

| **Model** | **n** | **AUC (95% CI)** | **Sensitivity (95% CI)** | **Specificity (95% CI)** |
|---|---|---|---|---|
| *Command Only* | | | | |
| Adjacency Eigenvalues | 3362 | 0.88 (0.83-0.93) | 0.89 (0.80-0.96) | 0.74 (0.70-0.77) |
| Laplacian Eigenvalues | 3362 | 0.88 (0.84-0.93) | 0.78 (0.67-0.89) | 0.86 (0.84-0.89) |
| Normalized Laplacian Eigenvalues | 3362 | 0.84 (0.78-0.90) | 0.87 (0.78-0.94) | 0.69 (0.65-0.72) |
| *Copy Only* | | | | |
| Adjacency Eigenvalues | 3362 | 0.87 (0.83-0.92) | 0.85 (0.76-0.94) | 0.78 (0.74-0.81) |
| Laplacian Eigenvalues | 3362 | 0.87 (0.82-0.92) | 0.89 (0.80-0.96) | 0.80 (0.76-0.83) |
| Normalized Laplacian Eigenvalues | 3362 | 0.88 (0.83-0.92) | 0.91 (0.81-0.98) | 0.74 (0.70-0.77) |
| *Command and Copy* | | | | |
| Adjacency Eigenvalues | 3362 | 0.86 (0.81-0.91) | 0.76 (0.65-0.87) | 0.84 (0.81-0.87) |
| Laplacian Eigenvalues | 3362 | 0.87 (0.83-0.92) | 0.91 (0.81-0.98) | 0.73 (0.69-0.76) |
| Normalized Laplacian Eigenvalues | 3362 | 0.84 (0.79-0.90) | 0.87 (0.78-0.96) | 0.70 (0.67-0.74) |

**Table S7:** Descriptive summaries of participant-level dCDT networks by cognitive status and test condition. Values are computed at the participant level and summarized as mean (SD) for continuous measures and n (%) for indicator measures. Percentages are calculated within cognitive group and test condition. Indicators include presence of isolated nodes and omission of clock components (e.g., missing digits or hands).

| | Command | | Copy | |
|---|---|---|---|---|
| *Measure* | Intact | Impaired | Intact | Impaired |
| At least one isolated node | 144 (4.70%) | 76 (28.10%) | 35 (1.10%) | 26 (9.60%) |
| Isolated nodes | 0.20 (1.32) | 1.32 (3.14) | 0.02 (0.33) | 0.38 (1.68) |
| Smallest edge weight | 0.43 (0.02) | 0.43 (0.03) | 0.43 (0.02) | 0.44 (0.04) |
| Largest edge weight | 0.96 (0.02) | 0.95 (0.04) | 0.96 (0.02) | 0.96 (0.02) |
| Mean edge weight | 0.63 (0.02) | 0.64 (0.03) | 0.63 (0.01) | 0.64 (0.03) |
| Smallest node degree (weighted) | 10.10 (2.27) | 7.63 (4.80) | 10.60 (1.16) | 9.73 (3.20) |
| Largest node degree (weighted) | 12.30 (0.95) | 11.70 (2.17) | 12.50 (0.33) | 12.40 (1.19) |
| Mean node degree (weighted) | 11.10 (1.20) | 10.20 (2.67) | 11.30 (0.40) | 11.10 (1.51) |
| Node degree variability | 0.64 (0.47) | 1.21 (1.14) | 0.56 (0.23) | 0.78 (0.76) |
| Diameter | 0.92 (0.02) | 0.92 (0.04) | 0.92 (0.02) | 0.92 (0.02) |
| Missing 1 digit | 51 (1.60%) | 23 (8.50%) | 5 (0.20%) | 8 (3.00%) |
| Missing 2 digit | 47 (1.50%) | 24 (8.90%) | 0 (0.00%) | 3 (1.10%) |
| Missing 3 digit | 35 (1.10%) | 19 (7.00%) | 3 (0.10%) | 5 (1.90%) |
| Missing 4 digit | 57 (1.80%) | 24 (8.90%) | 5 (0.20%) | 7 (2.60%) |
| Missing 5 digit | 52 (1.70%) | 27 (10.00%) | 9 (0.30%) | 9 (3.30%) |
| Missing 6 digit | 39 (1.30%) | 18 (6.70%) | 5 (0.20%) | 6 (2.20%) |
| Missing 7 digit | 58 (1.90%) | 29 (10.70%) | 4 (0.10%) | 7 (2.60%) |
| Missing 8 digit | 62 (2.00%) | 29 (10.70%) | 2 (0.10%) | 6 (2.20%) |
| Missing 9 digit | 33 (1.10%) | 24 (8.90%) | 4 (0.10%) | 7 (2.60%) |
| Missing 10 digit | 44 (1.40%) | 21 (7.80%) | 4 (0.10%) | 7 (2.60%) |
| Missing 11 digit | 42 (1.40%) | 24 (8.90%) | 8 (0.30%) | 4 (1.50%) |
| Missing 12 digit | 53 (1.70%) | 24 (8.90%) | 3 (0.10%) | 5 (1.90%) |
| Missing hour hand | 16 (0.50%) | 28 (10.40%) | 3 (0.10%) | 13 (4.80%) |
| Missing minute hand | 23 (0.70%) | 34 (12.60%) | 3 (0.10%) | 11 (4.10%) |
| Missing clock face | 5 (0.20%) | 2 (0.70%) | 2 (0.10%) | 1 (0.40%) |

### S3 Supplementary Figures

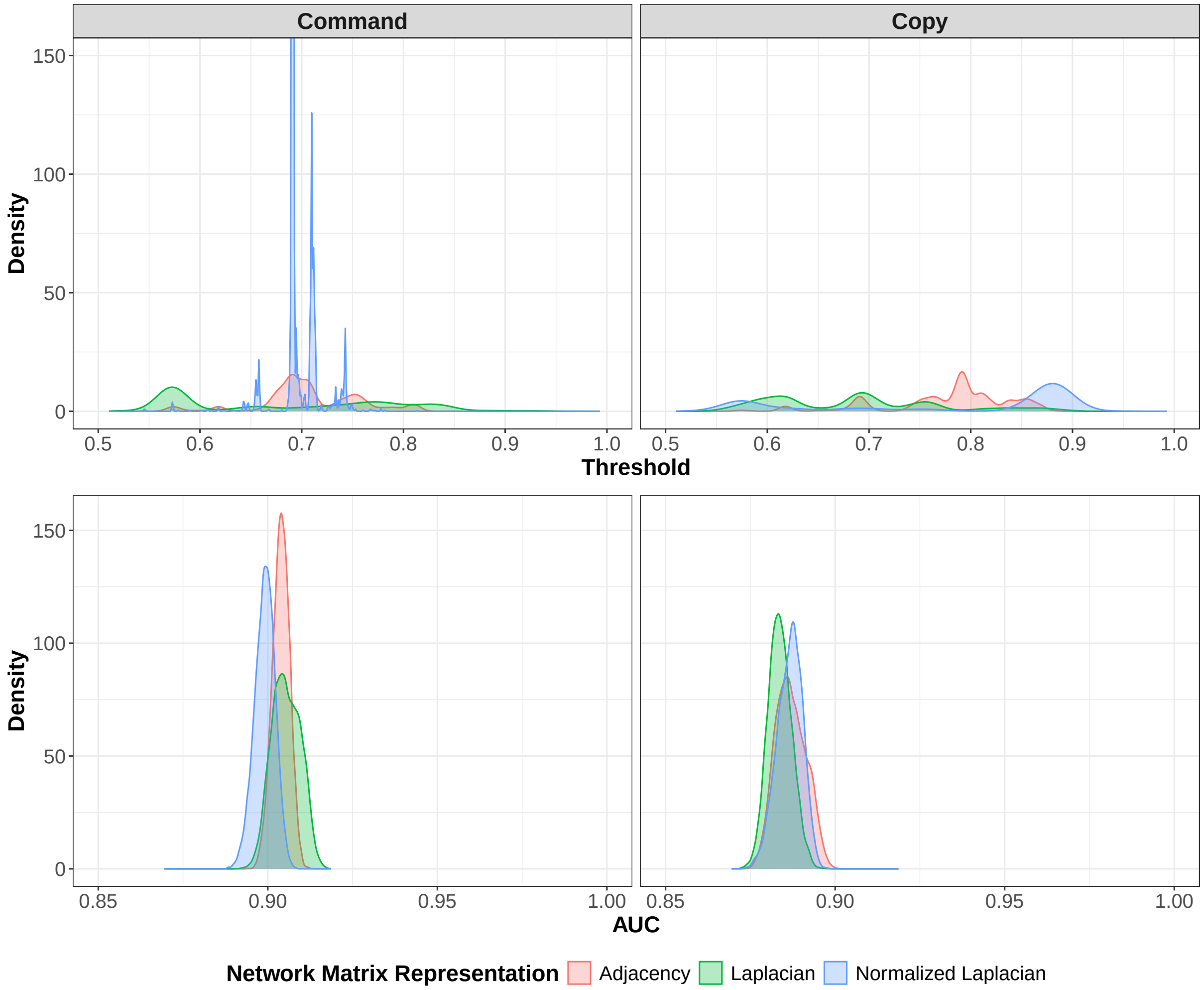


**Figure S1:** Distributions of empirically selected edge-weight threshold values across 10,000 resampled five-fold splits of the training data. For each split and each matrix representation (adjacency, Laplacian, normalized Laplacian), the threshold that maximized cross-validated AUC within the training data was identified separately for command and copy networks. The mode of each resulting distribution was selected as the final threshold for evaluation in the held-out validation set: 0.70 for command networks and 0.80 for copy networks, corresponding to the removal of edges with standardized Euclidean distances greater than 0.36 and 0.22, respectively. Density curves are truncated at 150 for visualization.
